# Age- and sex-specific total mortality impacts of the early weeks of the Covid-19 pandemic in England and Wales: Application of a Bayesian model ensemble to mortality statistics

**DOI:** 10.1101/2020.05.20.20107680

**Authors:** Vasilis Kontis, James E Bennett, Robbie M Parks, Theo Rashid, Jonathan Pearson-Stuttard, Perviz Asaria, Michel Guillot, Marta Blangiardo, Majid Ezzati

## Abstract

**Background:** The Covid-19 pandemic affects mortality directly through infection as well as through changes in the social, environmental and healthcare determinants of health. The impacts on mortality are likely to vary, in both magnitude and timing, by age and sex. Our aim was to estimate the total mortality impacts of the pandemic, by sex, age group and week.

**Methods:** We developed an ensemble of 16 Bayesian models that probabilistically estimate the weekly number of deaths that would be expected had the Covid-19 pandemic not occurred. The models account for seasonality of death rates, medium-long-term trends in death rates, the impact of temperature on death rates, association of death rates in each week on those in preceding week(s), and the impact of bank holidays. We used data from January 2010 through mid-February 2020 (i.e., week starting 15^th^ February 2020) to estimate the parameters of each model, which was then used to predict the number of deaths for subsequent weeks as estimates of death rates if the pandemic had not occurred. We subtracted these estimates from the actual reported number of deaths to measure the total mortality impact of the pandemic.

**Results:** In the week that began on 21^st^ March, the same week that a national lockdown was put in place, there was a >92% probability that there were more deaths in men and women aged ≥45 years than would occur in the absence of the pandemic; the probability was 100% from the subsequent week. Taken over the entire period from mid-February to 8^th^ May 2020, there were an estimated ~ 49,200 (44,700-53,300) or 43% (37-48) more deaths than would be expected had the pandemic not taken place. 22,900 (19,300-26,100) of these deaths were in females (40% (32-48) higher than if there had not been a pandemic), and 26,300 (23,800-28,700) in males (46% (40-52) higher). The largest number of excess deaths occurred among women aged >85 years (12,400; 9,300-15,300), followed by men aged >85 years (9,600; 7,800-11,300) and 75-84 years (9,000; 7,500-10,300).

The cause of death assigned to the majority (37,295) of these excess deaths was Covid-19. There was nonetheless a >99.99% probability that there has been an increase in deaths assigned to other causes in those aged ≥45 years. However, by the 8^th^ of May, the all-cause excess mortality had become virtually equal to deaths assigned to Covid-19, and non-Covid excess deaths had diminished to close to zero, or possibly become negative, in all age-sex groups.

**Interpretation:** The death toll of Covid-19 pandemic, in middle and older ages, is substantially larger than the number of deaths reported as a result of confirmed infection, and was visible in vital statistics when the national lockdown was put in place. When all-cause mortality is considered, the mortality impact of the pandemic on men and women is more similar than when comparing deaths assigned to Covid-19 as underlying cause of death.

## Introduction

The Covid-19 pandemic has led to tens of thousands of deaths among patients with confirmed infection in the UK. The pandemic has also profoundly changed the social, economic, environmental and healthcare determinants of morbidity and mortality. These changes are likely to impact public health beyond the deaths caused directly by infection through a number of routes^1^ including delayed disease prevention and procedures for acute and chronic medical care; loss of jobs and income; disruption of social networks; changes in crime and self-harm; changes in quantity and quality of food, and the use of tobacco, alcohol and other drugs; and changes in mobility and transport patterns with potential impacts on road traffic injuries and air pollution.^2^ How changes in these social, environmental and healthcare determinants impact mortality is likely to vary, in both magnitude and timing, by age and sex.

An understanding of the mortality impacts beyond deaths assigned to Covid-19 infection is needed to understand the overall public health impacts of the pandemic and control policies, titrate and adjust the response, and put in place mitigation mechanisms to minimize the adverse impacts as the pandemic and response continue beyond early weeks. We developed and applied methodology to quantify the weekly mortality impacts of the Covid-19 pandemic and associated responses by age group and sex in England and Wales. The methodology can also be used for comparable cross-country analysis on a real-time basis.

## Methods

### Data sources

We used data on the weekly number of deaths in England and Wales, by age group and sex released by the Office for National Statistics (ONS). Weekly death files include all deaths registered from Saturday through the subsequent Friday. They also include data on the number of deaths involving Covid-19, which are deaths with a mention of Covid-19 anywhere on the death certificate, including in combination with other health conditions. We used data from first week in January 2010 through the week starting on Saturday 2^nd^ May 2020 (ending on 8^th^ May 2020). We used data on mid-year population by age group and sex from the ONS. We calculated weekly population through interpolation, as done by the ONS for quarterly population.^3^ We obtained data on temperature from ERA5,^4^ which uses data from global in situ and satellite measurements to generate a worldwide meteorological dataset, with full space and time coverage over our analysis period. We used gridded estimates measured four times daily at a resolution of 30 km to generate weekly temperatures for each local authority district, which we weighted by local authority population to create national level summaries.

### Statistical methods

The total mortality impact of the Covid-19 pandemic should be calculated as the difference between the observed number of deaths and the number of deaths had the pandemic not occurred, which is not directly measurable. The most common approach to addressing this issue has been to use the average number of deaths over previous years, e.g., the most recent five years, for the corresponding week or month when the comparison is made.^5^ This approach however does not take into account changes in population size and age structure, nor long-and short-term trends in mortality, which are particularly pronounced for some age groups.^6,7^ Nor does this approach account for time-varying factors like temperature, that are largely external to the pandemic, but also affect death rates. In addition, bank holidays such as Easter that do not occur in the same week of the year affect number of deaths and death registration.

We developed an ensemble of 16 short-term Bayesian mortality projection models that each make an estimate of weekly death rates that would be expected if the Covid-19 pandemic had not occurred. We used multiple models because there is inherent uncertainty in the choice of model that best predicts death rates in the absence of pandemic. These models were formulated to incorporate features of weekly death rates as follows:

- First, death rates may have a medium-to-long-term trend.^6-8^ We developed two sets of models, one with no trend and one with a linear trend term over weekly deaths.
- Second, death rates have a seasonal pattern, which varies by age group and sex.^9-12^ We included weekly random intercepts for each week of the year. To account for the fact that seasonal patterns “repeat” (i.e., late December and early January are seasonally similar) we used a seasonal structure^13,14^ for the random intercepts. The seasonal structure allows the magnitude of the random intercepts to vary over time, and implicitly accounts for time-varying factors such as annual fluctuations in flu season.
- Third, death rates in each week may be related to those in preceding week(s). We formulated four sets of models to account for this relationship. The weekly random intercepts in these models had a first, second, fourth or eighth order autoregressive structure^13,14^ The higher order autoregressive models allow death rates in any given week to be informed by those in a progressively larger number of preceding weeks. Further, trends not picked up by the linear or seasonal terms would be captured by these autoregressive terms.
- Fourth, beyond having a seasonal pattern, death rates depend on temperature, and specifically on whether temperature is higher or lower than its long-term norm during a particular time of year.^15-20^ The effect of temperature on mortality varies throughout the year, and may be in opposite directions for different times of year. We used two sets of models, one without temperature and one with a weekly term for temperature anomaly, defined as deviation of weekly temperature from the local average weekly temperature over the entire analysis period. The coefficients of temperature anomalies were specified as a random effect with a random walk prior of order one, so that temperature effect is more similar in adjacent weeks. The random effect had a circular structure so that late December and early January are treated as adjacent.
- Fifth, reported death rates in weeks that contain or follow a holiday may be different from other weeks. We included effects (as fixed intercepts) for the week containing and the week after each of the following holidays: Christmas and/or boxing day; Good Friday; Easter Monday; New Year’s day; early-May bank holiday; spring bank holiday; and summer bank holiday.
- We also tested, but did not include, model terms for the weeks that coincided with a change to and from daylight saving time because the effect was negligible.

These choices led to an ensemble^21^ of 16 short-term Bayesian mortality projection models (2 trend options x 4 autoregressive options x 2 temperature options). We used the time-series of weekly reported deaths from January 2010 through mid-February 2020 (i.e., week starting 15^th^ February 2020) to estimate the parameters of each model, which was then used to predict death rates for subsequent weeks as estimates of the counterfactual death rates (i.e., if the pandemic had not occurred). For the projection period, we used recorded temperature so that our projections take into consideration actual temperature in 2020. This choice of training and prediction periods assumes that the number of deaths that are directly or indirectly related to the Covid-19 pandemic was negligible through mid-February 2020, which is about two weeks after the first confirmed case in the UK, but it allows for impacts to have appeared in subsequent weeks.

We used weakly informative log gamma priors on log precision with both shape and rate equal to 0.01. We tested the sensitivity of the results to the choice of prior through the use of penalized complexity priors and found that the results were similar. All models were fitted using integrated nested Laplace approximation (INLA),^22^ implemented in the R-INLA software (version 20.03). We took 1,000 draws from the posterior distribution of age-specific deaths under each model, and pooled the 16,000 draws to obtain the posterior distribution of age-specific deaths if the Covid-19 pandemic had not taken place. The reported credible intervals represent the 2.5^th^ and 97.5^th^ percentiles of the posterior distribution of the draws from the entire ensemble. This approach incorporates both the uncertainty of estimates from each model and the uncertainty in the choice of model.

We did all analyses separately by sex and age group (0-14 years, 15-44 years, 45-64 years, 65-74 years, 75-84 years and 85+ years) because death rates, and how they are impacted by the pandemic, vary by age group and sex. To obtain estimates across age groups and both sexes, we summed draws from age-sex-specific estimates. For the purpose of reporting, we rounded results on number of deaths that are ≥1000 to the nearest hundred to avoid giving a false sense of precision in the presence of uncertainty; results <1000 are rounded to the nearest ten.

We tested the performance of the projections from the model by withholding data for 11 weeks starting from mid-February (i.e., the same projection period as done for 2020) for an earlier year and used the preceding time-series of data to train the models. We then projected death rates for the weeks with withheld data, and evaluated how well the model ensemble projections reproduce the known-but-withheld death rates. We repeated this for three different years: 2017 (i.e. train model using data from January 2010 to mid-February 2017 and test for the subsequent 11 weeks), 2018 (i.e., train model using data from January 2010 to mid-February 2018 and test for the subsequent 11 weeks), and 2019 (i.e., train model using data from January 2010 to mid-February 2019 and test for the subsequent 11 weeks). We report the projection error (which measures systematic bias) and absolute forecast error (which measures any deviation from the data). Additionally, we report coverage of the projection uncertainty; if projected death rates and their uncertainties are well estimated, the estimated 95% credible intervals should cover 95% of the withheld data.

## Results

Weekly mortality varied substantially over time in all ages with evidence of seasonal pattern above 45 years of age (Figure 1). From 22^nd^ February through 20^th^ March 2020, the observed number of deaths in every age group was well within the credible interval of what was expected to have occurred if the Covid-19 pandemic had not taken place (Figures 1 and 2). In the week that began on 21^st^ March, the same week that a national lockdown was put in place, there was already a >92% probability that there were more deaths in both sexes and all age groups ≥45 years than would occur in the absence of the pandemic; the probability was 100% (i.e., every one of the 16,000 draws were positive) from the subsequent week (Figures 2 and 3). The same phenomenon occurred in those aged 15-44 years in the first week of April 2020.

**Figure 1.**
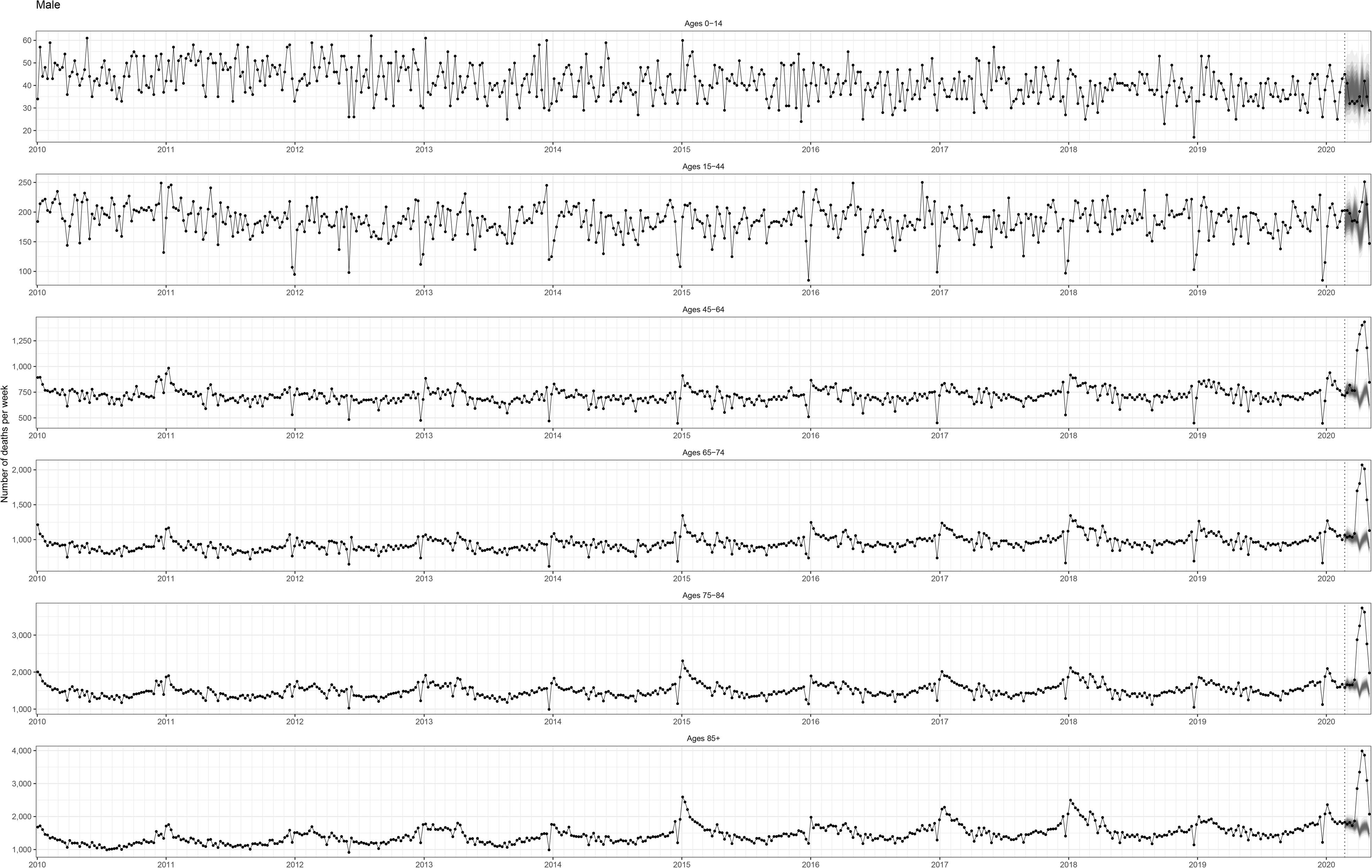
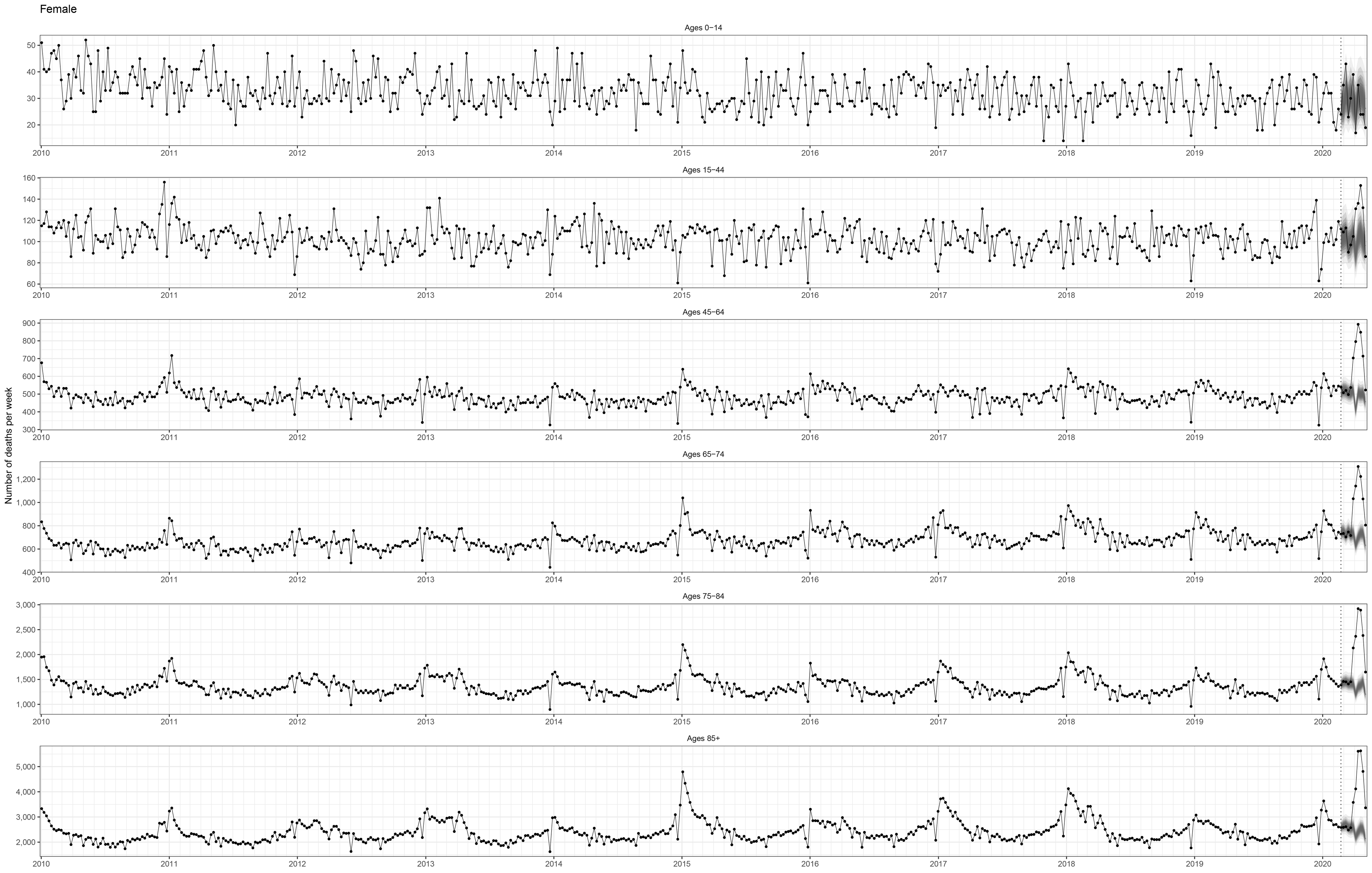
Weekly number of deaths by age group from January 2010 through 8^th^ May 2020. The black points show reported deaths. Each point is placed at the beginning of the week. The grey-shaded areas show the posterior distribution of how many deaths would have occurred from the week starting on 22^nd^ February 2020 (shown in the vertical dashed line) through the week starting on 2^nd^ May 2020 (ending on 8^th^ May 2020) had Covid-19 pandemic not taken place. The grey shading shows the levels of credible intervals around the median prediction, from 5% (dark grey) to 95% (light grey).

**Figure 2.**
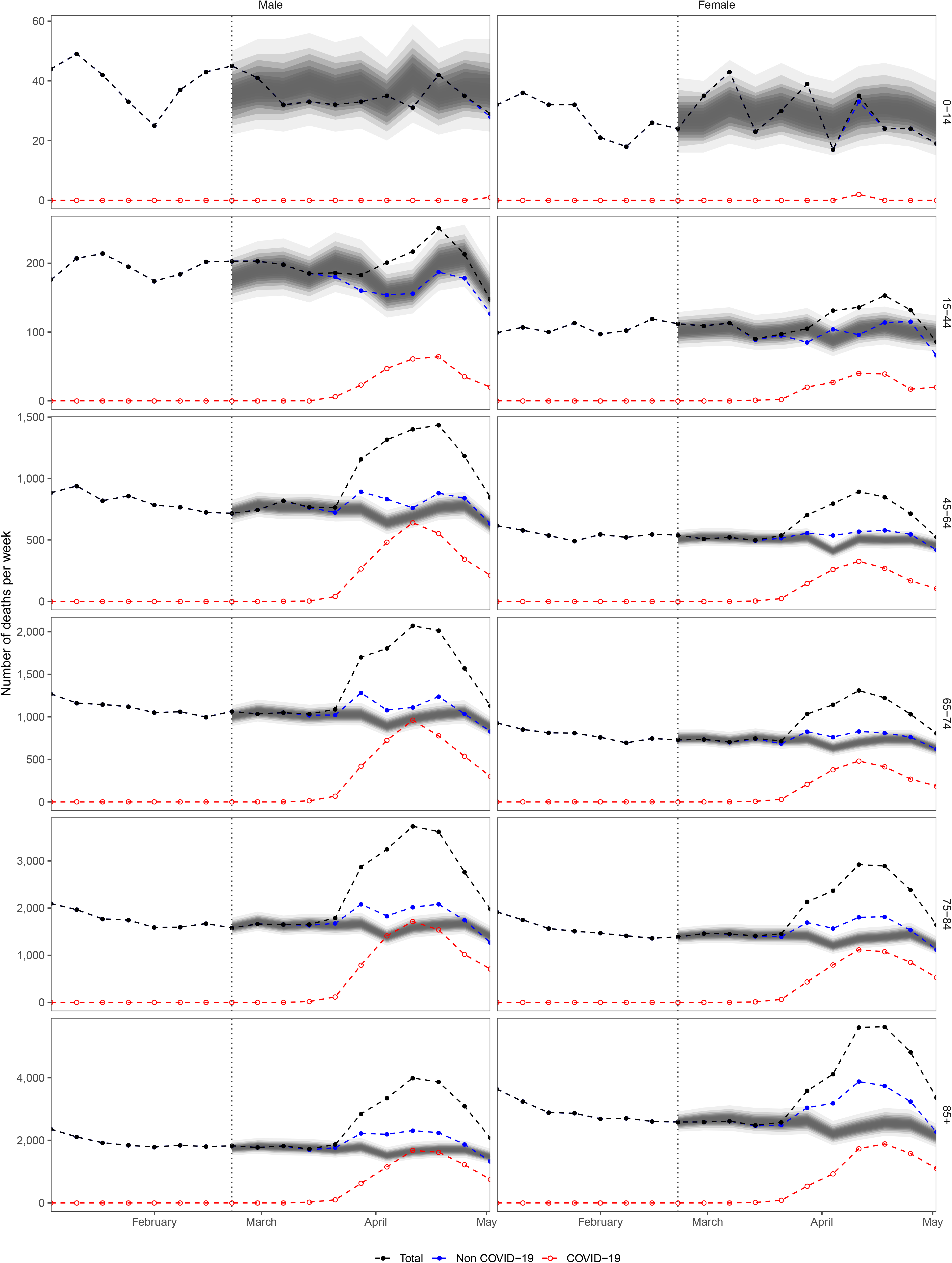
Weekly number of deaths by age group from January 2020 through 8^th^ May 2020. The points show reported deaths: black for total; red for Covid-19 and blue for other causes of death. Each point is placed at the beginning of the week. The grey-shaded areas show the predictions of how many deaths would have been expected from the week starting on 22^nd^ February 2020 (shown in the vertical dashed line) through the week starting on 2^nd^ May 2020 (ending on 8^th^ May 2020) had Covid-19 pandemic not taken place. The grey shading shows the levels of credible intervals around the median prediction, from 5% (dark grey) to 95% (light grey).

**Figure 3.**
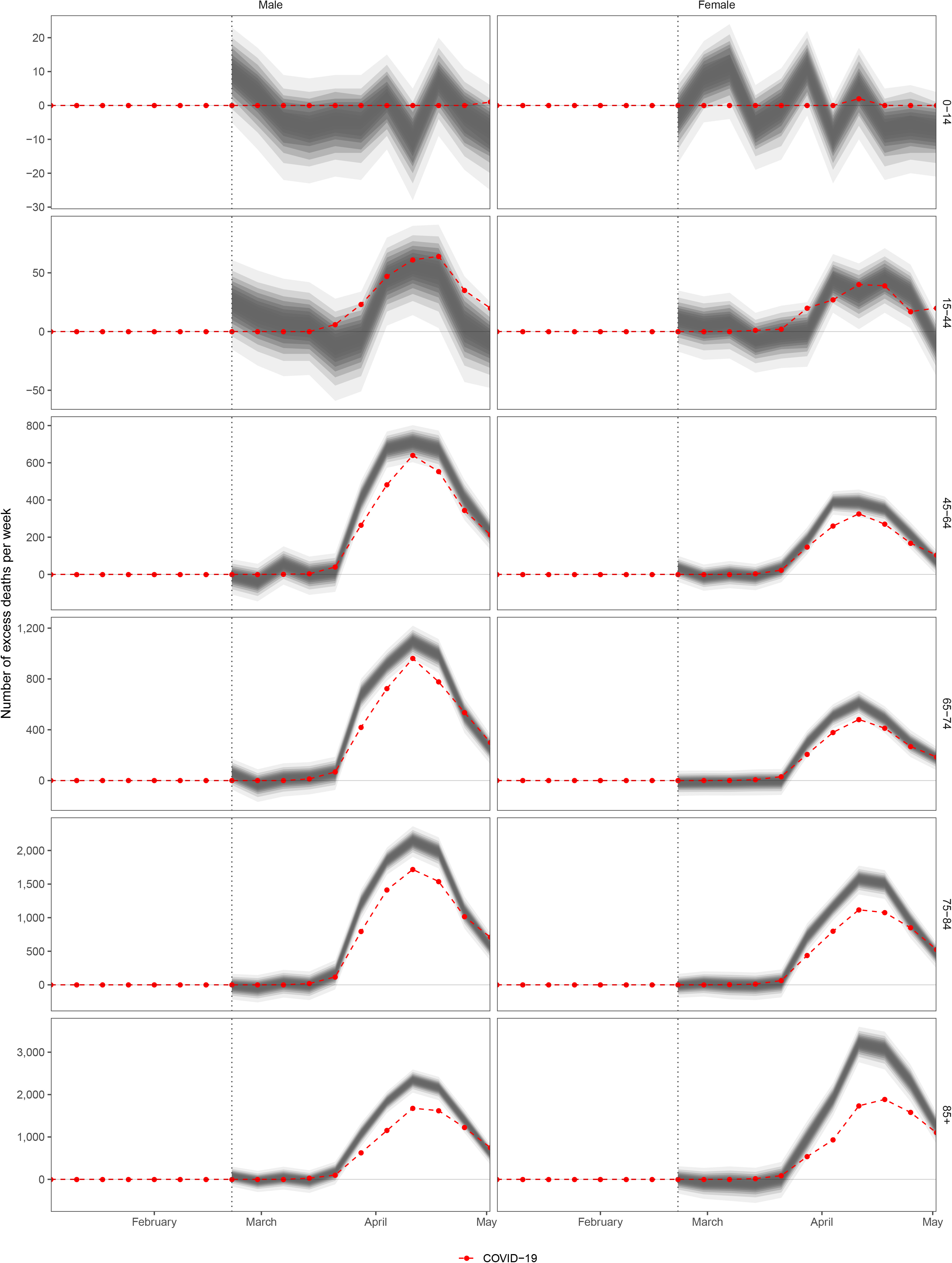
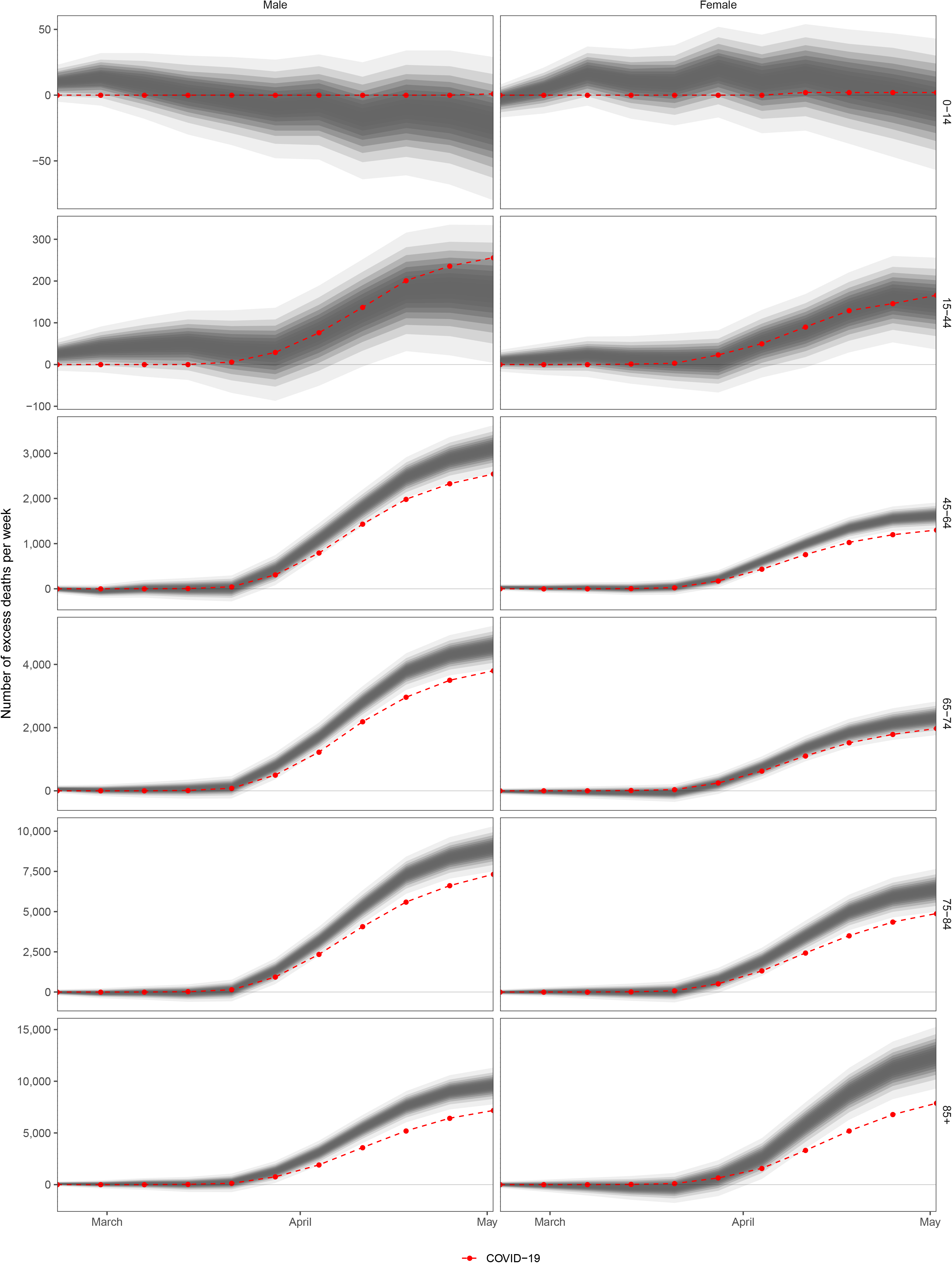
Estimated number of deaths due to Covid-19 pandemic. (A) weekly deaths and (B) cumulative deaths. The grey shading shows the levels of credible intervals around the median prediction, from 5% (dark grey) to 95% (light grey). The red points show the number of deaths assigned to Covid-19 as underlying cause of death. The difference between these points and the curves is excess non-Covid-19 deaths.

Taken over the entire period from mid-February to 8^th^ May 2020 and across all age groups, 164,373 deaths were registered in England and Wales. This represents an estimated ~ 49,200 (44,700-53,300) or 43% (37-48) more deaths than would be expected had the pandemic not taken place. 22,900 (19,300-26,100) of these deaths were in females (40% (32-48) higher than if there had not been a pandemic), and 26,300 (23,800-28,700) in males (46% (40-52) higher). The largest overall (i.e. from any cause) number of excess deaths occurred among women aged >85 years (12,400; 9,300-15,300), followed by men aged >85 years (9,600; 7,800-11,300) and 75-84 years (9,000; 7,500-10,300).

The cause of death assigned to the majority (37,295) of these excess deaths was Covid-19 (Figures 2 and 3). Nonetheless, there was a >99.99% probability that there has also been an increase in deaths assigned to other causes in those aged ≥45 years. The share of total excess deaths not assigned to Covid-19 was higher in those aged 75 years and older and was particularly high in 85+ year olds. Specifically, over the entire period from 22^nd^ February to 8^th^ May 2020, the share of non-Covid excess deaths was 18%, 17%, 18% and 25% in men aged 45-64, 65-74, 75-84 and 85+ years, respectively, and 20%, 15%, 23% and 37% in women aged 45-64, 65-74, 75-84 and 85+ years. The number of non-Covid excess deaths also increased with age, reaching 1,600 (180-3,000) and 1,500 (110-2,800) in men and women aged 75-84 years, respectively, and 2,400 (580-4,100) and 4,600 (1,400-7,400) in men and women aged 85+ years. However, by the 8^th^ of May, non-Covid excess deaths had diminished to close to zero or possibly become negative in all age-sex groups. In boys and young men aged 0-14 and 15-44 years, there may have been a short-term decline, lasting two weeks in mid-late March, in non-Covid deaths but the probability that deaths were lower than would be expected without the pandemic in different weeks was <80%. Similarly, in the first week of May, deaths in boys/men and girls/women aged <45 years may have dropped below what would be expected without the pandemic with posterior probabilities ranging from 69% to 89%.

The results of model validation (Table 2) show that the estimates of how many deaths would be expected in different weeks had the pandemic not occurred had mean projection errors <6% in all age-sex groups. The mean absolute error was also <9% in all age-sex groups except in those aged 0-14 years where the number of deaths is small. 95% coverage, which measures how well the posterior distributions of projected deaths coincide with withheld data was >93% for all age and sex groups which shows that the posterior distribution is well-estimated.

**Table 1.** Number of deaths by age group and sex. Numbers in brackets show 95% credible intervals. We report the number of deaths registered by the ONS without rounding. We rounded the estimated number of excess deaths that are ≥1000 to the nearest one hundred and those <1000 to the nearest ten.

| Age group (years) | Number of deaths | Deaths expected if no pandemic | Excess deaths due to pandemic (all causes) | Deaths assigned to Covid-19 | Excess deaths from other causes |
| --- | --- | --- | --- | --- | --- |
| Male |  |  |  |  |  |
| 0-14 | 388 | 410<br>(360-470) | -20<br>(-80-30) | 1 | -20<br>(-80-30) |
| 15-44 | 2,187 | 2,000<br>(1,900-2,200) | 180<br>(0-330) | 256 | -80<br>(-250-80) |
| 45-64 | 11,148 | 8,000<br>(7,500-8,600) | 3,100<br>(2,600-3,600) | 2,541 | 570<br>(10-1,100) |
| 65-74 | 15,560 | 11,000<br>(10,300-11,700) | 4,600<br>(3,900-5,200) | 3,799 | 750<br>(60-1,400) |
| 75-84 | 26,557 | 17,600<br>(16,200-19,100) | 9,000<br>(7,500-10,300) | 7,326 | 1,600<br>(180-3,000) |
| 85+ | 28,233 | 18,600<br>(16,900-20,500) | 9,600<br>(7,800-11,300) | 7,176 | 2,400<br>(580-4,100) |
| Total | 84,073 | 57,700<br>(55,300-60,300) | 26,300<br>(23,800-28,700) | 21,099 | 5,200<br>(2,700-7,600) |
| Female |  |  |  |  |  |
| 0-14 | 313 | 320<br>(270-370) | -0<br>(-60-40) | 2 | -10<br>(-60-40) |
| 15-44 | 1,264 | 1,100<br>(1,000-1,200) | 150<br>(40-260) | 166 | -20<br>(-130-90) |
| 45-64 | 7,081 | 5,500<br>(5,200-5,800) | 1,600<br>(1,300-1,900) | 1,301 | 320<br>(20-610) |
| 65-74 | 10,174 | 7,900<br>(7,300-8,400) | 2,300<br>(1,800-2,800) | 1,973 | 340<br>(-210-860) |
| 75-84 | 21,509 | 15,100<br>(13,800-16,500) | 6,400<br>(5,000-7,700) | 4,881 | 1,500<br>(110-2,800) |
| 85+ | 39,959 | 27,500<br>(24,700-30,700) | 12,400<br>(9,300-15,300) | 7,873 | 4,600<br>(1,400-7,400) |
| Total | 80,300 | 57,400<br>(54,200-61,000) | 22,900<br>(19,300-26,100) | 16,196 | 6,700<br>(3,100-9,900) |

**Table 2.** Results of the validation projections from the Bayesian model ensemble. Each number represents the average across three validation runs, using data through mid-February 2017, 2018 and 2019 respectively, and forecasting 11 weeks ahead.

| Age group<br>(years) | Projection error<br>(relative projection error) |  | Absolute projection error<br>(relative absolute projection error) |  | Percent covered by 95%<br>credible interval |  |
| --- | --- | --- | --- | --- | --- | --- |
|  | M | F | M | F | M | F |
| 0-14 | 20 (5.17%) | -5 (-1.03%) | 66 (15.95%) | 67 (20.04%) | 96.97% | 93.94% |
| 15-44 | 14 (0.88%) | 1 (0.08%) | 179 (8.80%) | 97 (8.77%) | 96.97% | 100.00% |
| 45-64 | -45 (-0.50%) | 27 (0.52%) | 335 (4.02%) | 280 (5.07%) | 100.00% | 100.00% |
| 65-74 | 193 (1.75%) | 312 (3.99%) | 595 (5.29%) | 530 (6.62%) | 93.94% | 93.94% |
| 75-84 | 583 (3.40%) | 673 (4.57%) | 1,146 (6.48%) | 1,120 (7.28%) | 93.94% | 96.97% |
| 85+ | 563 (3.28%) | 1,002<br>(3.78%) | 1,356 (7.39%) | 2,520 (8.64%) | 96.97% | 93.94% |

## Discussion

We applied a robust probabilistic method to coherently and consistently estimate the total death toll of Covid-19 pandemic from the 22^nd^ February to 8^th^ May. At this stage, the Covid-19 pandemic was responsible for over 49,000 excess deaths in England and Wales. We also found that when all-cause mortality is considered, the mortality impact of the pandemic on men (~46% increase in deaths) and women (~40% increase in deaths) is more similar than when comparing deaths assigned to Covid-19. Deaths that were not assigned to Covid-19 made up 24% of all excess deaths, but had diminished in most age-sex groups by the first week of May. We also rule out, with virtual certainty (posterior probability >99.99%), the hypothesis that the death toll of the pandemic may be smaller than direct Covid-19 deaths either because some of those dying of Covid-19 would have died in this same time period from other underlying conditions even if the pandemic had not occurred,^23,24^. Further, although children and young adults may have experienced shot-term declines in deaths, taken across all age groups any “positive” impacts of the lockdown (e.g., reductions in deaths due to air pollution or traffic injuries) are dwarfed by the negative impacts of the epidemic.

Our overall estimates are similar to those reported by the ONS^25^ and the Financial Times^5^ but our findings reveal important details on excess deaths by age group and sex which these sources do not. EuroMoMo does not report country-specific excess deaths and hence could not be compared with our results.

The main strength of our work is the systematic use of time-series data from 2010 to early 2020 to estimate how many deaths would be expected in the absence of pandemic. By modelling death rates, rather than simply the number of deaths as is done in most other analyses, we account for changes in population size and age structure. The models incorporated important features of mortality, including seasonality of death rates, how mortality in one week may depend on previous week(s) and the seasonally-variable role of ambient temperature. The use of a modelling framework, as we have done, allowed us to make estimates by age group and sex, and, as data become available, will allow doing so for specific causes of death and subnational geographies which would, because of smaller numbers, not be possible for other methods. We used an ensemble of models which typically leads to more robust projections and represent both the uncertainty associated with each individual model and that of model choice.^21^ Finally, this framework, specifically the inclusion of seasonality and ambient temperature, is well suited for more robust estimation and standardised comparisons of excess deaths across countries on a real-time basis.

The main limitation of our work is that we did not have data on underlying cause of death beyond the distinction between Covid-19 and non-Covid deaths. Having a breakdown of deaths by underlying cause will help develop cause-specific models and understand which causes have exceeded or fallen below the levels expected. We also could not access age-specific data by region because the ONS only releases aggregate numbers for regions. Nor did we have data on total mortality by socio-demographic status to understand inequalities in the impacts of the pandemic beyond deaths assigned to Covid-19 as the underlying cause of death. Releasing these data will allow more granular analysis of the impacts of the pandemic, which can in turn inform resource allocation and a more targeted approach to mitigating both the direct and indirect effects of Covid-19, now and for future waves of the pandemic. Further, weekly mortality files from the ONS cover deaths registered in any given week. These include some deaths from prior weeks and leave out some deaths in the reporting week.^26^ However, the approach is consistent over time and does not affect year to year comparisons, including for 2020 as the lags in registration of deaths assigned to Covid-19 seem to be the same as those from other causes.^27^

It is likely that some of the apparently non-Covid excess deaths are due to undetected Covid-19 infections.^28^ An example of such deaths are the likely Covid-19 deaths in care homes.^29,30^ Other such deaths may be those who called the NHS helpline or the ambulance service, were advised to self-isolate because their symptoms were not deemed sufficiently severe to be admitted, and died at home.^31,32^ That the share of excess deaths from non-Covid causes became smaller over time may be because with increasing awareness of, and attention to, clinical symptoms more of such deaths are assigned to Covid-19 as the underlying cause. It is also possible that many excess deaths have been caused, and may continue to do so in coming months, by the pandemic due to changes in personal and family economic and employment circumstances, and in healthcare provision, access and utilisation, as evidenced by reductions in A&E attendance and procedures for a diverse range of acute and chronic conditions.^33-41^ While the official position on this has been to encourage people to seek care, the situation is more complex: for some accessing care becomes more restricted because their family members are infected and cannot continue supporting them. Others, typically those in limited and marginalised housing and employment, may not do so in fear of losing their livelihood.

The large death toll of the pandemic, from deaths assigned to Covid-19 as well as other causes, together with the fact that excess death toll was already happening when a national lockdown was announced indicate that the cessation of community contact tracing in early March, hesitation in putting a lockdown in place earlier, and sub-optimal identification and management of those with complex conditions in the community at increased risk from both the virus and effects of lockdown, is likely to have contributed to the substantial excess deaths. Minimising these impacts requires a coherent strategy that supresses the epidemic and strengthens the social safety net and healthcare provision, in facilities as well as in the community and at home, together with transparent communication to encourage resumption of care seeking.

## Data Availability

Data are obtained from the Office for National Statistics (ONS) and ERA5.

## Acknowledgement

We thank Giulia Mangiameli for help with background materials and references.

## Author contributions

All authors contributed to study design. VK and JEB developed and tested statistical methods with input from other authors. VK, RMP, TR and JEB accessed, harmonised and analysed data. VK conducted analysis and prepared results. ME wrote the first draft of the paper and other authors contributed to the paper.

## References

1. Douglas M, Katikireddi SV, Taulbut M, McKee M, McCartney G. Mitigating the wider health effects of covid-19 pandemic response. BMJ 2020; 369: m1557.

2. European Environment Agency. Air quality and COVID-19. 2020. https://www.eea.europa.eu/themes/air/air-quality-and-covid19/air-quality-and-covid19 (accessed 6 Apr 2020.

3. Office for National Statistics. Quarterly mortality report, England: October to December 2019 and year-end review 2019.

4. ECMWF. ERA5: Reanalysis Datasets 2019.

5. Giles C. Excess UK deaths in Covid-19 pandemic top 50,000 Financial Times 2020.

6. Bennett JE, Li G, Foreman K, et al. The future of life expectancy and life expectancy inequalities in England and Wales: Bayesian spatiotemporal forecasting. Lancet 2015; 386(9989): 163–70.

7. Bennett JE, Pearson-Stuttard J, Kontis V, Capewell S, Wolfe I, Ezzati M. Contributions of diseases and injuries to widening life expectancy inequalities in England from 2001 to 2016: a population-based analysis of vital registration data. Lancet Public Health 2018; 3(12): e586-e97.

8. Kontis V, Mathers CD, Rehm J, et al. Contribution of six risk factors to achieving the 25×25 non-communicable disease mortality reduction target: a modelling study. Lancet 2014; 384(9941): 427–37.

9. Parks RM, Bennett JE, Foreman KJ, Toumi R, Ezzati M. National and regional seasonal dynamics of all-cause and cause-specific mortality in the USA from 1980 to 2016. e Life 2018; 7.

10. Feinstein CA. Seasonality of Deaths in the U.S. by Age and Cause. Demogr Res 2002; 6(17): 471–88.

11. Fowler T, Southgate RJ, Waite T, et al. Excess Winter Deaths in Europe: a multi-country descriptive analysis. European Journal of Public Health 2014; 25(2): 339–45.

12. McKee CM. Deaths in winter: Can Britain learn from Europe? European journal of epidemiology 1989; 5(2): 178–82.

13. Hyndman RJ, Athanasopoulos G. Forecasting: principles and practice, 2nd edition. Melbourne, Australia: OTexts; 2018.

14. Congdon P. Applied Bayesian Modelling John Wiley & Sons; 2003.

15. Basu R. High ambient temperature and mortality: a review of epidemiologic studies from 2001 to 2008. Environ Health 2009; 8: 40.

16. Basu R, Samet JM. Relation between elevated ambient temperature and mortality: a review of the epidemiologic evidence. Epidemiol Rev 2002; 24(2): 190–202.

17. Gasparrini A, Guo Y, Hashizume M, et al. Mortality risk attributable to high and low ambient temperature: a multicountry observational study. Lancet 2015; 386(9991): 369–75.

18. Song X, Wang S, Hu Y, et al. Impact of ambient temperature on morbidity and mortality: An overview of reviews. Sci Total Environ 2017; 586: 241–54.

19. Bennett JE, Blangiardo M, Fecht D, Elliott P, Ezzati M. Vulnerability to the mortality effects of warm temperature in the districts of England and Wales. Nature Clim Change 2014; 4(4): 269–73.

20. Parks RM, Bennett JE, Tamura-Wicks H, et al. Anomalously warm temperatures are associated with increased injury deaths. Nat Med 2020; 26(1): 65–70.

21. Hoeting JA, Madigan D, Raftery AE, Volinsky CT. Bayesian model averaging: a tutorial. Statistical Science 1999; 14: 382–401.

22. Rue H, Martino S, Chopin N. Approximate Bayesian inference for latent Gaussian models using integrated nested Laplace approximations (with discussion). Journal of the Royal Statistical Society, Series B 2009; 71(2): 319–92.

23. Knapton S. Two thirds of coronavirus victims may have died this year anyway, government adviser says The Telegraph. 2020 25 March 2020.

24. Coker R. Harvesting’ is a terrible word but it’s what has happened in Britain’s care homes. The Guardian. 2020 8 May 2020.

25. Office for National Statistics. Deaths registered weekly in England and Wales, provisional: week ending 8 May 2020; 2020.

26. Office for National Statistics. Comparison of weekly death occurrences in England and Wales: up to week ending 8 May 2020 2020.

27. Office for National Statistics. Deaths involving COVID-19, England and Wales: deaths occurring in April 2020; 2020.

28. Fuller T, Baker M. Coronavirus Death in California Came Weeks Before First Known U.S. Death. The New York Times. 2020 22 Apr 2020.

29. Perraudin F. Coronavirus involved in quarter of care home residents’ deaths in England and Wales The Guardian. 2020 15 May 2020.

30. The Health Foundation. Care homes have seen the biggest increase in deaths since the start of the outbreak, 2020.

31. Laville S. London woman dies of suspected Covid-19 after being told she was ‘not priority’. The Guardian. 2020 25 Mar 2020.

32. Nagesh A. The Uber driver evicted from home and left to die of coronavirus. BBC news. 2020 28 April 2020.

33. Campbell D, Bannock C. Coronavirus crisis could lead to 18,000 more cancer deaths, experts warn The Guardian. 2020 29 April 2020.

34. NHS. Cancelled Elective Operations Data. 2020. https://www.england.nhs.uk/statistics/statistical-work-areas/cancelled-elective-operations/cancelled-ops-data/.

35. Sample I. More than 2m operations cancelled as NHS fights Covid-19. The Guardian. 2020 26 Apr 2020.

36. Hiom S. How coronavirus is impacting cancer services in the UK. In: UK CR, editor. Science blog: Cancer Research UK; 2020.

37. Campbell D. Record drop in A&E attendance in England ‘a ticking timebomb’, say doctors The Guardian. 2020 14 May 2020.

38. Bernstein L, Sellers FS. Patients with heart attacks, strokes and even appendicitis vanish from hospitals. The Washington Post. 2020 19 Apr 2020.

39. Pell MB, Lesser B. COVID’s Other Casualties. Reuters Investigates. 2020 3 April 2020.

40. Kansagra AP, Goyal MS, Hamilton S, Albers GW. Collateral Effect of Covid-19 on Stroke Evaluation in the United States. The New England journal of medicine 2020.

41. Gaudino M, Chikwe J, Hameed I, Robinson NB, Fremes SE, Ruel M. Response of Cardiac Surgery Units to COVID-19: An Internationally-Based Quantitative Survey. Circulation 2020.

